# Olfactory Bulb and Amygdala Gene Expression Changes in Subjects Dying with COVID-19

**DOI:** 10.1101/2021.09.12.21263291

**Authors:** Ignazio S. Piras, Matthew J. Huentelman, Jessica E. Walker, Richard Arce, Michael J. Glass, Daisy Vargas, Lucia I. Sue, Anthony J. Intorcia, Courtney M. Nelson, Katsuko E. Suszczewicz, Claryssa L. Borja, Marc Desforges, Michael Deture, Dennis W. Dickson, Thomas G. Beach, Geidy E. Serrano

**Affiliations:** Translational Genomics Research Institute, Neurogenomics Division; Banner Sun Health Research Institute, Sun City, AZ; Centre Hospitalier Universitaire Sainte-Justine, Laboratory of Virology, Montreal, Canada; Mayo Clinic College of Medicine, Mayo Clinic Florida, Jacksonville, FL

**Author notes:** Correspondence: Geidy E. Serrano, Banner Sun Health Research Institute, 10515 West Santa Fe Drive, Sun City, AZ 85351.

## Abstract

In this study we conducted RNA sequencing on two brain regions (olfactory bulb and amygdala) from subjects who died from COVID-19 or who died of other causes. We found several-fold more transcriptional changes in the olfactory bulb than in the amygdala, consistent with our own work and that of others indicating that the olfactory bulb may be the initial and most common brain region infected. To some extent our results converge with pseudotime analysis towards common processes shared between the brain regions, possibly induced by the systemic immune reaction following SARS-CoV-2 infection. Changes in amygdala emphasized upregulation of interferon-related neuroinflammation genes, as well as downregulation of synaptic and other neuronal genes, and may represent the substrate of reported acute and subacute COVID-19 neurological effects. Additionally, and only in olfactory bulb, we observed an increase in angiogenesis and platelet activation genes, possibly associated with microvascular damages induced by neuroinflammation. Through coexpression analysis we identified two key genes (*CAMK2B* for the synaptic neuronal network and *COL1A2* for the angiogenesis/platelet network) that might be interesting potential targets to reverse the effects induced by SARS-CoV-2 infection. Finally, in olfactory bulb we detected an upregulation of olfactory and taste genes, possibly as a compensatory response to functional deafferentation caused by viral entry into primary olfactory sensory neurons. In conclusion, we were able to identify transcriptional profiles and key genes involved in neuroinflammation, neuronal reaction and olfaction induced by direct CNS infection and/or the systemic immune response to SARS-CoV-2 infection.

## INTRODUCTION

The coronavirus SARS-CoV-2 is primarily associated with severe respiratory disease, termed COVID-19 disease, but there have also been numerous clinical reports of an accompanying broad range of neurological signs, symptoms and syndromes, affecting up to 36% of patients ^1-10^, although it is not clear whether these are due to direct viral brain effects or to systemic reactions to critical illness, including coagulopathy, sepsis, autoimmune mechanisms or multiorgan failure ^11^.

There is abundant evidence that coronaviruses are able to invade the CNS, and two previous pandemic coronaviruses, SARS-CoV and MERS-CoV, are documented to have caused similar human syndromes as has have been observed with COVID-19 ^12-38^. Two human coronaviruses, strains 229E and OC43, were detected by RT-PCR, Northern hybridization and in-situ hybridization in 44% and 23%, respectively, of 90 human brains obtained from multiple brain banks throughout Europe, the UK and North America ^32^, suggesting that, once brain invasion occurs, coronaviruses may persist for long periods of time in the human brain.

There have been more than 20 published studies ^11,39-58^ that used RT-PCR methods to interrogate SARS-CoV-2 genomic presence in postmortem brain tissue, and although results have differed between studies, the overall conclusion that most of these investigators have reached is that SARS-CoV-2 brain invasion occurs in only a relatively small fraction of those that have died of COVID-19 and that Ct values and viral copy numbers are generally low, perhaps only representing residual genomic fragments. It is apparent that differing PCR protocols also differ in their sensitivity and specificity ^58-60^ and therefore the true prevalence of viral brain entry and proliferation is still uncertain. The detection of subgenomic SARS-CoV-2 sequences has been suggested to prove that *in situ* viral replication has taken place. However, even in COVID-19 lung tissue, in one study only a subset of cases were found positive for these ^40^ and another study found that subgenomic fragments in the throat were only present in the first week of infection ^61^. One group has speculated that viral brain infection may be a “transient phenomenon” ^62^. It may therefore prove difficult to unequivocally identify the postmortem CNS presence of viable, reproducing SARS-CoV-2.

Of possible CNS entry points, both clinical and autopsy evidence has converged on the olfactory bulb, with its immediate neural connection to the olfactory sensory epithelium in the nasopharynx ^54,58,63-65^. Whole or partial loss of the sense of smell is present in 60% of COVID-19 patients ^64,66,67^. The olfactory mucosa strongly expresses angiotensin converting enzyme-2 (ACE2) and neuropilin-1, probable cellular access cofactors for SARS-CoV-2 ^15,20,68-71^. While there have been almost two dozen reports using PCR to localize SARS-CoV-2 RNA in postmortem COVID-19 brain tissue, only a few of these have assayed more than a very few brain regions. One study that assayed multiple brain regions found positive olfactory bulb amplification in 8/15 (53%) while all other brain regions except midbrain were negative ^58^. This result parallels that of our own ^72^ study (with follow up results in preparation) in which olfactory bulb was PCR-positive in 8/20 cases (40%) while of a total of 16 brain regions assayed, the next most-commonly-positive area was amygdala, with only 2 positive cases (10%). However, aside from olfactory bulb, the prevalence and copy numbers of PCR-detected SCV2 viral genome in autopsied COVID-19 brains seems insufficient to account for the much more prevalent neurological signs and symptoms.

Also puzzling is the relative lack of microscopic brain pathology in the great majority of autopsied COVID-19 cases. Although cases of meningitis and/or encephalitis have occurred ^46,51,52,73-78^, elements of the classical neuropathology of viral CNS infections ^79,80^, including lymphocytic leptomeningitis and encephalitis, microglial nodules, perivascular lymphocytic cuffing, focal demyelination and viral inclusions, have most often been absent. The lesions that have more been frequently reported, such as those with acute and subacute ischemic and/or hemorrhagic features ^45,51,53,54,56,73,78,81-88^, are common in unselected autopsy series. As almost all studies have not included appropriate non-COVID-19 control cases, it is difficult to know what lesions are specifically due to, or are more common in, COVID-19 brains. We found that acute and subacute ischemia or infarction was present in the brains of 14% of 691 consecutive pre-COVID-19 autopsies with or without concurrent autopsy-proven pneumonia, while the rate of acute brain hemorrhage was 1.4% and 2.0% for those with or without acute pneumonia, respectively ^89^. In comparison, in reviews of COVID-19 publications, reported clinically-determined rates of acute brain infarction range from 0.5% to 20% while rates of acute brain hemorrhage range from 0.13% to 2%^4,5,11,44,45,47,50,52-54,56,57,73,78,82,84-88,90-103^.

With direct viral brain damage seemingly uncommon in COVID-19, it appears to be more likely that its neurological manifestations are mostly due to systemic reactions that are common to critical illnesses, and may therefore be lacking specific histopathological correlates, but may be reflected by functional changes. Gene expression is a functional change that is sensitive to both local and systemic influences and may occur without accompanying alterations in cellular structure. Cell infection by SARS-CoV-2 may represent a direct stress, which will induce changes in cellular transcriptome. HCoV-OC43 infection of neuronal cells has been shown to induce the unfolded protein response. Changes in CNS gene expression could account for much of the neurological spectrum of COVID-19. We therefore used RNA sequencing technology to assess regional brain gene expression in postmortem brains of subjects that died with COVID-19, focusing on the olfactory bulb and the closely-connected amygdala.

## MATERIALS AND METHODS

### Human Subjects and Characterization

The human subjects were derived from Banner Sun Health Research Institute (BSHRI) in Sun City, Arizona (n =10), and from the Mayo Clinic in Jacksonville, Florida (n = 10). Clinical, neuropathological and RT-PCR results, including brain regional results, are described in our prior publication ^72^ and are briefly re-summarized in Table 1. All COVID-19 subjects had positive clinical diagnostic test results for SARS-CoV-2 and all were considered to have died in 2020 as a result of severe COVID-19. Since our prior publication ^72^, we have performed additional SARS-CoV-2 PCR assays, using alternate protocols (manuscript in preparation), on frozen samples of amygdala and olfactory bulb and these detected SARS-CoV-2 RNA in olfactory bulb from 6 additional cases, giving a new total of 8/20 SARS-CoV-2-positive cases in olfactory bulb. For amygdala these new protocols detected one additional case, giving a new total of 2 SARS-CoV-2-positive cases in amygdala.

**Table 1.** Presence of dementia, parkinsonism and neuropathological diagnoses in study subjects, with SCV2 PCR results in olfactory bulb and amygdala for the 20 COVID-19 subjects. B1-10 = BSHRI-derived COVID-19 cases; M1-10 = Mayo Clinic-derived COVID-19 cases; M9 was deleted as this subject was later found not to have had COVID-19; C1-20 = BSHRI-derived non-COVID19 control cases. SCV2 = PCR-positive for SARS-CoV-2. Abbreviations for major neuropathologically-diagnosed conditions: AD = Alzheimer’s disease dementia; FTLD-TDP = frontotemporal lobar degeneration with TDP-43 proteinopathy; HS = hippocampal sclerosis; MSA = multiple system atrophy; None = age-consistent changes only; PD = Parkinson’s disease; PSP = progressive supranuclear palsy; VaD = vascular dementia. Pneumonia in control cases refers to autopsy-confirmed non-COVID-19 pneumonia (pre-COVID-19 era); Control cases in this table were not assayed for SCV2; non-COVID-19 control cases were all negative for SCV2 in our prior publication^72^.

| Case | Age | Sex | Olf. Bulb<br>SCV2 | Amygdala<br>SCV2 | Clinical Diagnoses | Neuropathological<br>Diagnoses |
| --- | --- | --- | --- | --- | --- | --- |
| B1 | 90s | M | - | - | COVID-19;<br>Dementia | AD; VaD; HS; PSP |
| B2 | 70s | F | - | - | COVID-19;<br>Dementia | PD; AD |
| B3 | 70s | M | - | - | COVID-19;<br>No dementia or<br>parkinsonism | None |
| B4 | 80s | M | + | + | COVID-19;<br>No dementia or<br>parkinsonism | None |
| B5 | 90s | M | + | - | COVID-19;<br>Dementia, parkinsonism | AD |
| B6 | 70s | M | - | - | COVID-19;<br>Dementia, parkinsonism | AD; PSP |
| B7 | 70s | F | + | - | COVID-19;<br>No dementia or<br>parkinsonism | None |
| B8 | 80s | M | - | - | COVID-19;<br>Dementia | AD; MSA |
| B9 | 70s | M | + | - | COVID-19;<br>No dementia or<br>parkinsonism | None |
| B10 | 60s | F | - | + | COVID-19;<br>No dementia or<br>parkinsonism | None |
| M1 | 70s | F | - | - | COVID-19;<br>Dementia | AD; VaD |
| M2 | 80s | F | - | - | COVID-19;<br>Dementia | AD |
| M3 | 70s | F | - | - | COVID-19;<br>Dementia | DLB; AD |
| M4 | 80s | M | + | - | COVID-19;<br>Parkinsonism | PSP |
| M5 | 70s | F | - | - | Dementia,<br>parkinsonism | PSP |
| M6 | 60s | M | - | - | COVID-19;<br>Dementia | AD |
| M7 | 70s | F | - | - | COVID-19;<br>Dementia | FTLD-TDP; VaD |
| M8 | 70s | M | + | - | COVID-19;<br>Dementia, parkinsonism | VaD; AD |
| M10 | 30s | M | + | - | COVID-19;<br>No dementia or<br>parkinsonism | None |
| M11 | 90s | F | + | - | COVID-19;<br>Dementia | DLB |
| C1 | 70s | M | N/A | N/A | Pneumonia;<br>No dementia or<br>parkinsonism | None |
| C2 | 80s | M | N/A | N/A | Pneumonia;<br>No dementia or<br>parkinsonism | None |
| C3 | 70s | F | N/A | N/A | No pneumonia;<br>No dementia or<br>parkinsonism | None |
| C4 | 90s | M | N/A | N/A | Pneumonia;<br>No dementia or<br>parkinsonism | None |
| C5 | 80s | F | N/A | N/A | No pneumonia;<br>No dementia or<br>parkinsonism | None |
| C6 | 90s | F | N/A | N/A | No pneumonia;<br>No dementia or<br>parkinsonism | None |
| C7 | 100s | M | N/A | N/A | Pneumonia;<br>No dementia or<br>parkinsonism | None |
| C8 | 70s | F | N/A | N/A | No pneumonia;<br>No dementia or<br>parkinsonism | None |
| C9 | 80s | M | N/A | N/A | Pneumonia;<br>Dementia | AD |
| C10 | 70s | M | N/A | N/A | No pneumonia;<br>No dementia or<br>parkinsonism | None |
| C11 | 70s | M | N/A | N/A | Pneumonia;<br>Dementia; Parkinsonism | AD; PD |
| C12 | 70s | M | N/A | N/A | Pneumonia;<br>Dementia; Parkinsonism | AD; PD |
| C13 | 80s | F | N/A | N/A | Pneumonia;<br>Dementia; Parkinsonism | AD; PD |
| C14 | 80s | M | N/A | N/A | Pneumonia;<br>Dementia | AD |
| C15 | 90s | M | N/A | N/A | No pneumonia;<br>Dementia | AD; VaD |
| C16 | 80s | M | N/A | N/A | Pneumonia;<br>Dementia; Parkinsonism | PD |
| C17 | 90s | F | N/A | N/A | No pneumonia;<br>Dementia | AD |
| C18 | 80s | F | N/A | N/A | No pneumonia;<br>No dementia or<br>parkinsonism | None |
| C19 | 90s | F | N/A | N/A | No pneumonia;<br>Dementia; Parkinsonism | AD; PSP; HS;<br>FTLD-TDP; VaD |
| C20 | 80s | F | N/A | N/A | No pneumonia;<br>Dementia; Parkinsonism | AD |

Olfactory bulb and amygdala brain tissue from additional human subjects (n = 20) without COVID-19 (who died prior to the COVID-19 pandemic) were used as a gene expression control group. Their basic characteristics are also listed in Table 1. The mean ages of the COVID-19 and non-Covid-19 control groups are 77.5 (SD 12.9) and 83.9 (SD 8.9) respectively (ns); each group has 9 females and 11 males. The control group includes equal numbers of subjects with and without autopsy-proven non-COVID-19 pneumonia. The COVID-19 group differs from the non-COVID-19 control group in the fraction with neuropathologically-diagnosed neurodegenerative conditions (6/20 vs 10/20, respectively) but this was not significant on a Fisher Exact test (p = 0.33).

### Gene Expression

RNA was extracted from 20 mg of frozen tissue using Qiagen RNeasy Plus mini kits (Qiagen, cat #74134) following the manufacturer’s instructions. The RNA Integrity Number (RIN) was assessed using Agilent’s RNA 6000 Nano Kit (Agilent, cat # 5067-1511) on the Bioanalyzer 2100. The mean RIN was greater than 8.0. Sequencing libraries were prepared with 100 ng of total RNA using Illumina’s Stranded Total RNA Prep Ligation with Ribo-Zero Plus (Illumina, Inc. cat # 20040529) following the manufacturer’s protocol. The final library was sequenced by 2 × 100bp paired-end sequencing on an Illumina NovaSeq6000.

After sequencing, FASTQs files were aligned to the Human Reference Genome HG38 using STAR ^105^, and summarized at the gene level with HTSeq. Quality controls were conducted using MultiQC software ^106^ and Principal Component Analysis (PCA). Samples with RIN < 4, total number of reads < 20M, and uniquely mapped reads < 70% were excluded from the downstream analysis. Genes with a total count of less than 10 were excluded. Normalization was performed using DEseq2. Differential gene expression was calculated between the COVID-19 samples and the controls using DEseq2, adjusting for age at death, sex, brain tissue source (Banner or Mayo Clinic) and neuropathologically-determined presence or absence of a diagnostic level of a major neurodegenerative disease. The presence or absence of non-COVID-19 pneumonia in control cases was also considered but preliminary analyses (not shown) indicated this was not a significance influence. Comparison of olfactory bulb gene expression in COVID-19 cases with or without PCR-detected SARS-CoV-2 showed very few differences in gene expression so subsequent comparisons involving olfactory bulb did not subdivide the COVID-19 cases on this basis (preliminary data, not shown). Genes with a False Discovery Rate (FDR) smaller than 0.05 were considered differentially expressed genes (DEGs). Pathway analysis was conducted on the DEGs using a hypergeometric statistic referencing the REACTOME database as implemented in the *clusterProfiler* R package.

Lists of cell specific genes were generated from a brain single-nucleus RNA sequencing dataset from the DLPFC ^107^ that included Alzheimer’s Disease cases (AD) and non-demented controls (ND) (total: n = 48). Data were downloaded from the Accelerated Medicine Partnership-AD portal (AMP-AD; accession number syn18485175). The filtered dataset consisted of a total of 70,634 droplet-based single-nuclei and 17,352 genes from: astrocytes (Ast), endothelial cells (End), excitatory neurons (Ex), inhibitory neurons (In), microglia (Mic), oligodendrocytes (Oli), oligodendrocyte precursor cells (Opc), and pericytes (Per). After excluding the AD samples, the data matrix was imported into Seurat ^108^ and normalized using the function “NormalizeData” with the option “LogNormalize”, using a scale factor of 10,000. Then, each gene was assigned to a cell class modeling a linear regression where the expression levels were the dependent variable and the cell type the predictors, adjusting for sex, age, and postmortem interval. A transcript was assigned to a cell type when: 1) the adjusted p-values (FDR) was p < 0.05, and 2) the regression coefficient of the enriched cell type had a ratio > 1.81 with the second enriched cell type. The cutoff was established after testing all of coefficient ratios from 1 to 5 until the variation of the number of unclassified genes stabilized to under 0.5%. We obtained a list of 5,641 cell-specific genes, which were used as gene sets to run enrichment analysis on the differentially expressed genes. The analysis was conducted using the R function *enrichment* (R package *bc3net*) adjusting the results for multiple testing with the FDR method.

We conducted WGCNA analysis to detect relevant coexpression networks associated with the disease ^109^. The count table was normalized with the *voom* method ^110^, and adjusted for the confounding factors as in the differential expression analysis using the function *removeBatchEffect* as implemented in *limma* ^111^. Genes with less than 5 total cpm counts were removed from the dataset. Finally, we retained the 50% of the most variable genes after computing the Median Absolute Deviation (MAD) with the goal of excluding genes with low variability that contribute noise in the coexpression networks. We computed soft-thresholding power (β), using the *pickSoftThreshold* function. Then, we plotted the values against the scale-free fit index and selected the lowest power for which the scale-free topology fit index curve flattens out upon reaching a r^2^ = 0.900 ^112^. We generated a signed coexpression network and identified the resulting clusters using the function *blockwiseModules* with the following parameters: *TOMtype*: “signed”, *minimum module size* = 30, *mergeCutHeight* = 0.30, *deepSplit* = 2; reassign threshold = 1.0^-06^, and *pamRespectsDendro* = “TRUE”. We computed the eigengene values for each individual and module by singular value decomposition (SVD) ^113^. The eigengenes were compared by module between COVID-19 positive and controls using a linear model as implemented in *limma*, adjusting the p-values for multiple testing by accounting for the number of modules using the FDR method. Covariates were not included in the model since we used the adjusted expression matrix. We conducted Gene Ontology (GO) enrichment analysis on the differentially expressed modules associated with COVID-19 status by the means of the R-package *enRichment*, using as background the intersection of given genes and genes present in GO. P-values of the GO enrichment analysis were adjusted using the Bonferroni method. The enrichment for genes specifically expressed in certain cell types was conducted using the above-described cell specific markers by means of a hypergeometric test as implemented in the R function *enrichment*. Finally, the top hub genes in the coexpression modules were identified using the function *chooseTopHubInEachModule* as implemented in the WGCNA package.

### Pseudotime analysis

Pseudotime analytical approaches are machine learning based algorithms capable of extracting latent temporal information from cross-sectional studies ^114^. First, the dataset was normalized and filtered as described for the WGCNA analysis. Then, pseudotime trajectories were generated using the *phenoPath* method modeling the brain region as a prior factor of interest ^114^. Finally, we correlated the gene expression to the pseudotime trajectories using the biweight midcorrelation to detect genes associated with the estimated disease progression ^109^ adjusting the results with the FDR method. Pathway analysis was conducted using the genes significantly correlated with pseudotime by means of the *clusterProfiler* R package.

## RESULTS

### Differential expression

After removing low quality samples, the final sample size was 38 for amygdala (AMY) (COVID-19+ = 18; CTL = 20) and 40 for olfactory bulb (OBT) (COVID-19+ = 20; CTL = 20). The remaining samples had a total of 5,003.6 million (M) of reads (range: 20.1M – 234.6M), with an average percentage of uniquely mapped reads of 82.8% (range: 72.8% - 90.3%). After differential expression analysis we obtained 1,283 DEGs in AMY (576 upregulated and 707 downregulated) (**Fig. 1A**). In the OBT samples we obtained 5,405 DEGs (2,546 upregulated and 2,859) dowregulated) (**Fig 1B**). *IFI6, INSRR* and *ITGA6* were the most significant genes in AMY, whereas *UBQLN2, CLCNA* and *MAPK1* were the most significant genes in OBT. The top 9 genes for the two regions are reported in **Figure 2A** and **2B**, whereas the complete list of DEGs is reported in **Table S1** and **S2**. Pathway analysis revealed differential processes associated with AMY and OBT. In AMY we observed an enrichment of immune pathways (including interferon signaling and the toll like receptor cascade), mostly due to upregulated genes. Across downregulated genes we observed an enrichment of synaptic and neuronal pathways (**Figure 3; Table S3**). In OBT we observed an enrichment of RNA metabolism and synaptic pathways. However, the top pathway enriched in upregulated genes was Olfactory signaling and collagen pathways. Finally, synaptic pathways were enriched within downregulated genes (**Figure 3; Table S4**).

**Figure 1.**
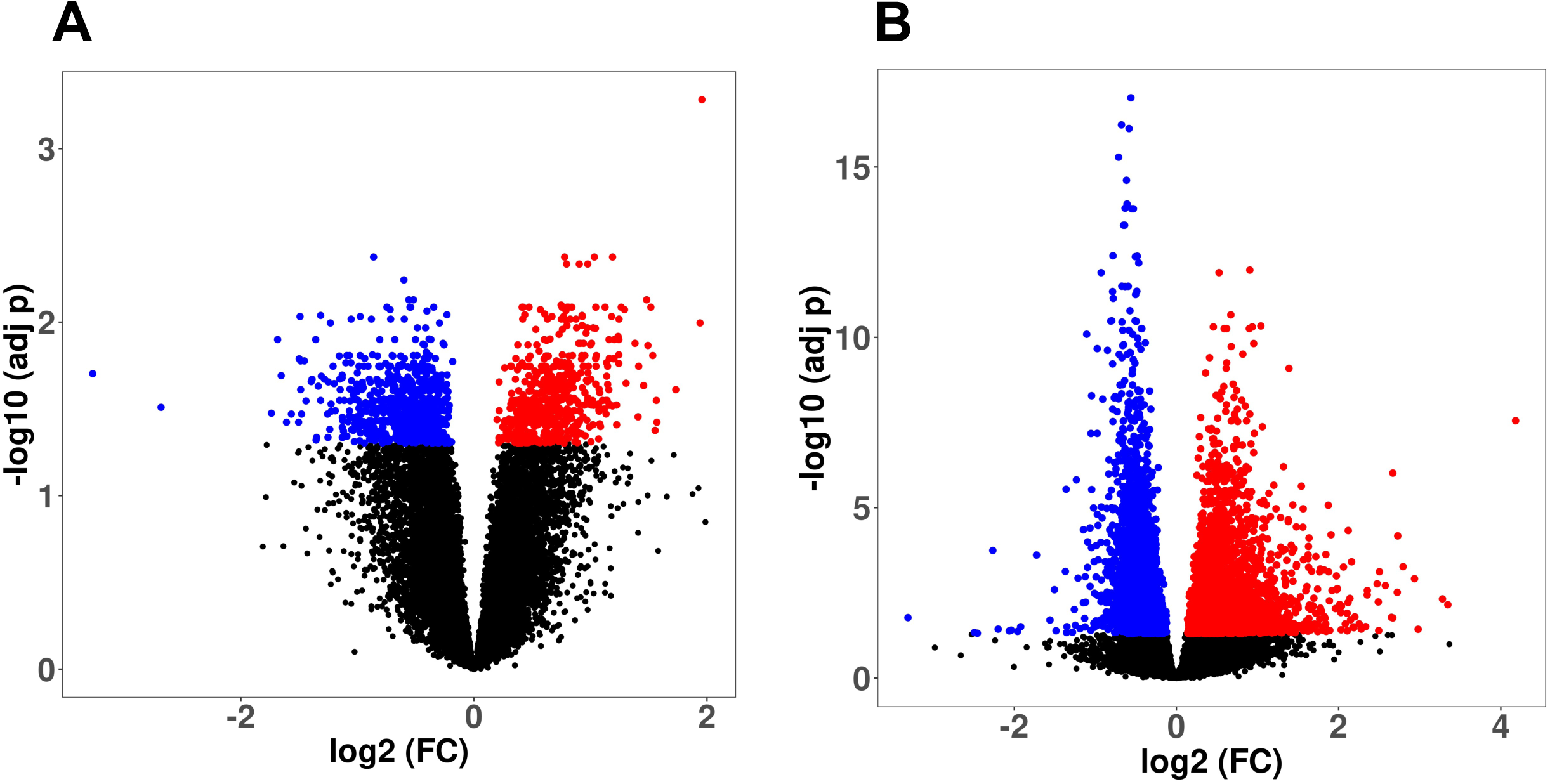
Volcano plot showing the differential expression analysis results after comparing Covid+ vs CTL in Amygdala (A) and Olfactory Bulb (B). Genes in blue and red were downregulated and upregulated in Covid+ patients (adj-p < 0.05).

**Figure 2.**
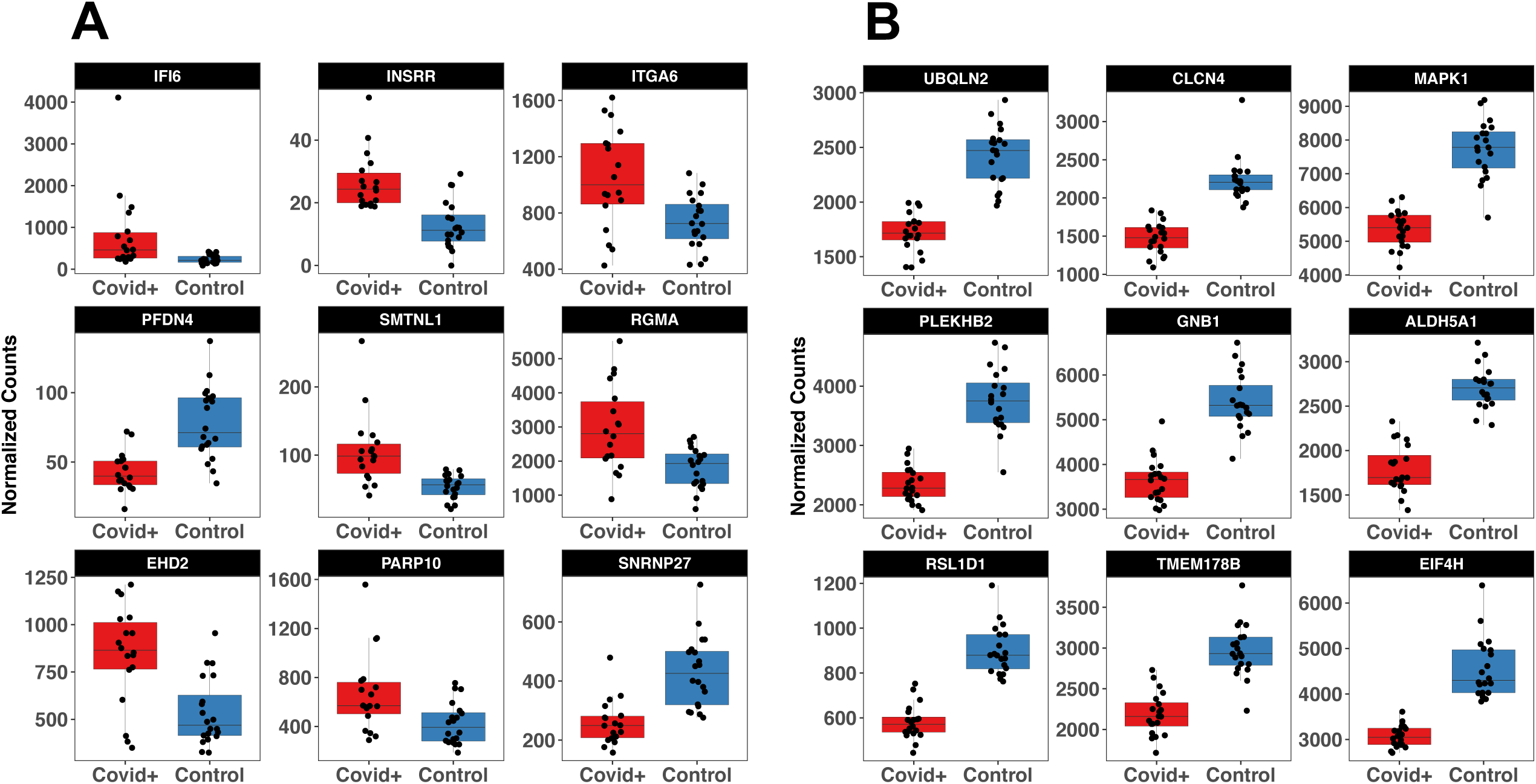
Top 9 genes differentially expressed in amygdala (A) and olfactory bulb (B)

**Figure 3.**
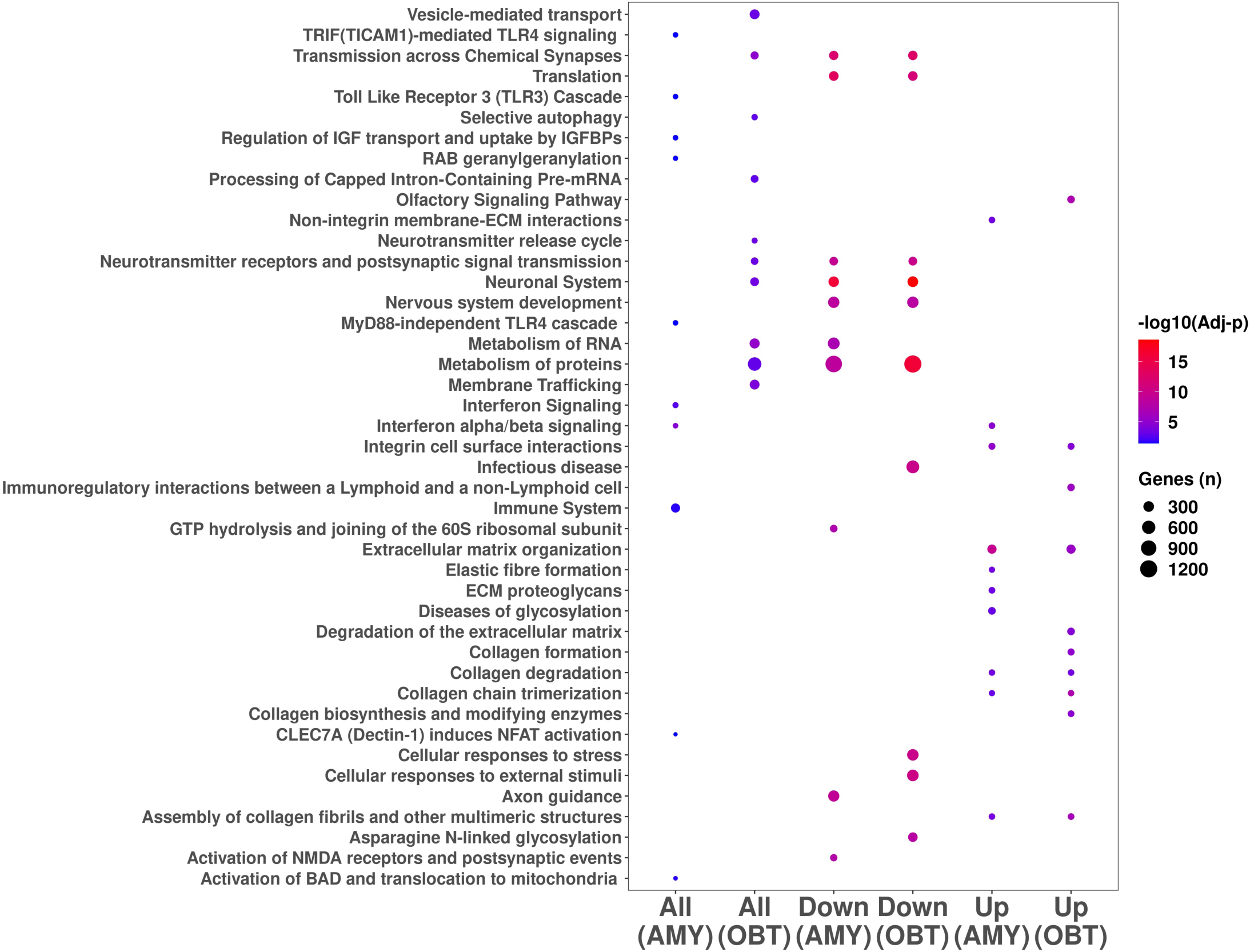
Top 10 pathways enriched in amygdala and olfactory bulb All: all DEGs; Up: DEGs upregulated in Covid+ patients; Down: DEGs downregulated in Covid+ patients

Cell specific gene enrichment across DEGs revealed astrocyte (Ast) and endothelial (End) specific genes were significantly over-represented in AMY. We also analyzed the enrichment results for upregulated and downregulated DEGs separately, detecting Ast, End, Per, Mic and Opc overrepresented in upregulated genes in AMY. Additionally, Ex and In were significantly enriched in the downregulated gene set in AMY. In OBT, when considering all the DEGs we did not detect any significant cell-specific enrichment. However, we detected a significant enrichment of End, Per, and Mic across upregulated genes, and enrichment of both Ex and In in the downregulated genes (**Figure 4**).

**Figure 4.**
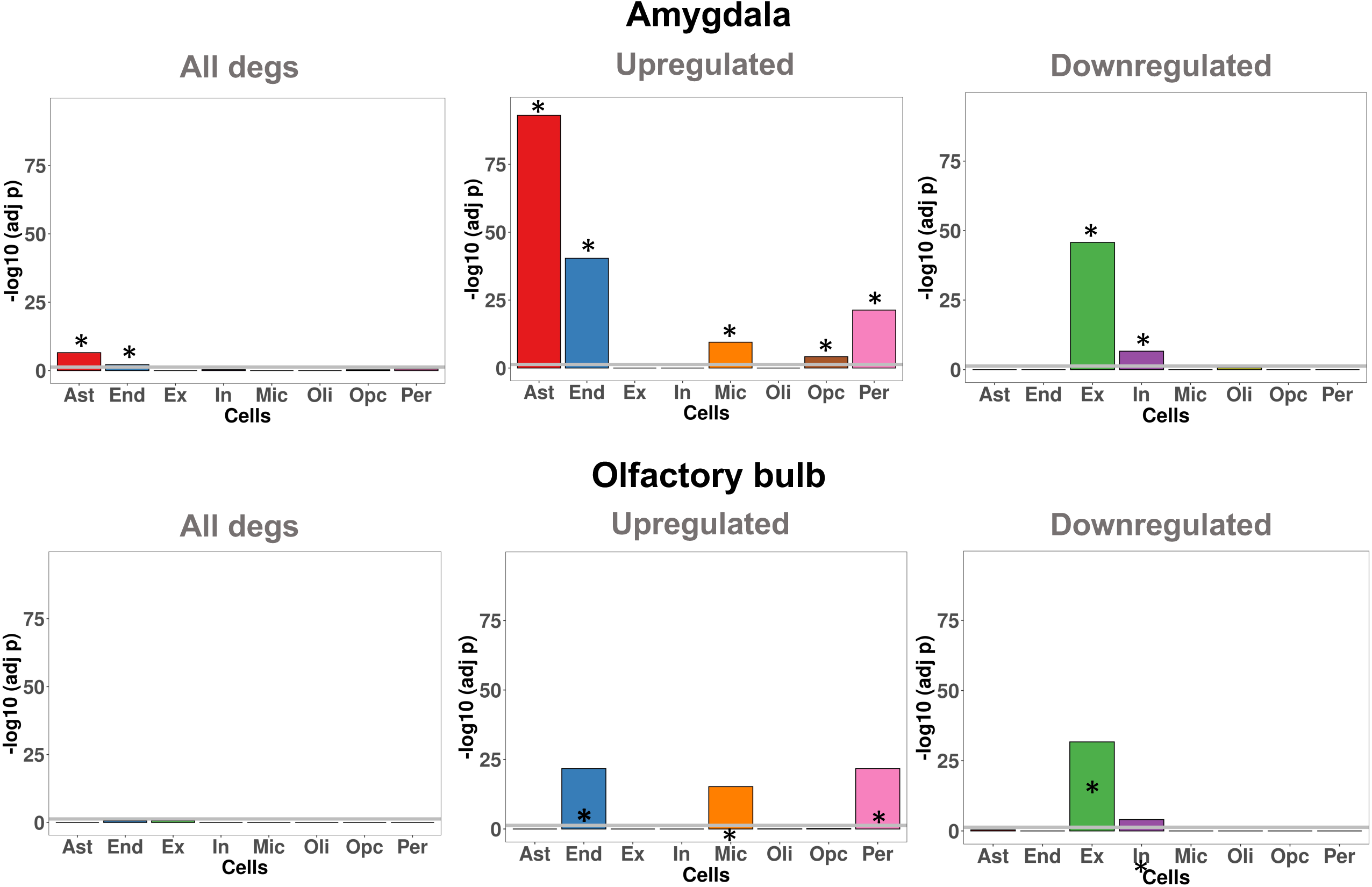
Cell specific genes enrichment in among DEGs in amygdala and olfactory bulb. Gene classes significantly enriched are indicate with the asterisk (FDR < 0.05)

### WGCNA analysis

We obtained a total of six coexpression modules (**Figure S1**) in amygdala with the number of genes in each module ranging from 204 (red module) to 3,049 (turquoise). After eigengene extraction and differential analysis, no modules were differentially expressed between COVID-19+ and CTL in AMY (**Figure S1**). In OBT we obtained 15 modules with the number of genes in each module ranging from 39 (midnightblue) to 1,904 (turquoise). Interestingly, 11 modules were differentially expressed between COVID-19+ and CTL, with the top modules being black (downregulated in COVID-19+) and red (upregulated) (both: FDR = 1.68^-11^) (**Figure 5**). Relationships between these modules are reported in **Figure S2**. We conducted Gene Ontology (GO) and cell enrichment analysis on these eleven modules and found GO enriched classes in eight of the modules. The top two modules (black and red) were enriched for ribosome/RNA metabolism and cilium/taste, respectively. Other modules were enriched for development/angiogenesis (green, upregulated, and enriched for End and Per), immune system (tan, upregulated), and synaptic signaling (green-yellow and turquoise, both downregulated and enriched for neuronal cell genes) (**Table S5**). Interestingly, the two synaptic modules were not related to each other (**Figure S2**) despite a similar functional enrichment. The summary of the WGCNA analysis, including modules, GO and cell enrichment, and a list of the hub genes is reported in **Table 2**.

**Figure 5.**
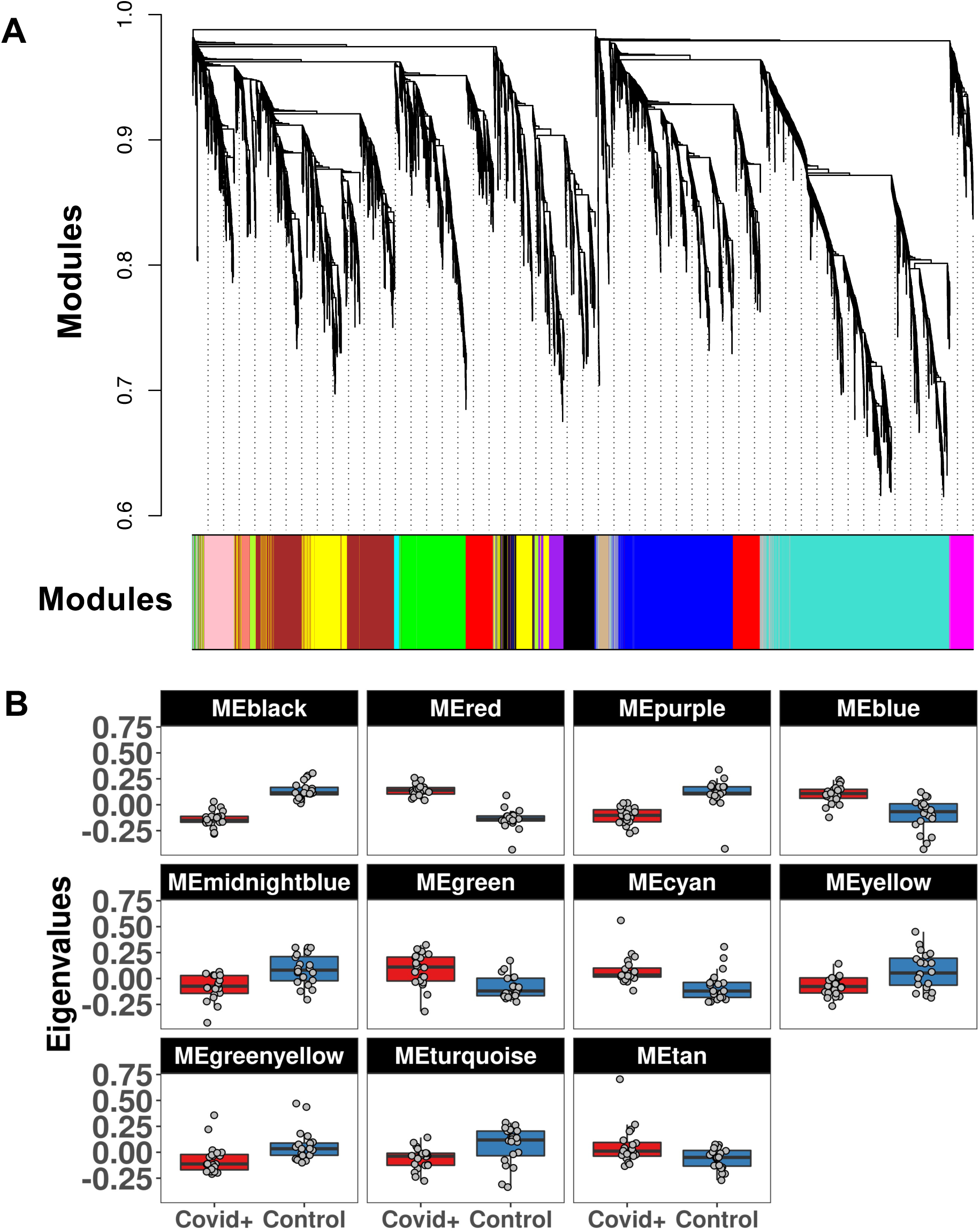
WGCNA analysis in olfactory bulb. We detected 15 coexpression modules (A) including 11 differentially expressed between Covid+ and Controls (B)

**Table 2.** Summary of the WGCNA analysis, including number of genes, differential analysis between Covid+ and controls, enrichment of pathways and cell specific genes, and hub genes for each co-expression network.

| Module | Genes (n) | Direction<br>(Covid+ vs CTL) | Adj-p | Cell | Pathways | Hub Gene |
| --- | --- | --- | --- | --- | --- | --- |
| Black | 459 | DOWN | 1.7E-11 | - | Ribosome/RNA metabolism | *CLSTN1* |
| Red | 587 | UP | 1.7E-11 | - | Cilium/Taste | *DMTF1* |
| Purple | 223 | DOWN | 3.5E-06 | - | Mitochondrion | *PJA2* |
| Blue | 1,248 | UP | 3.4E-05 | - | - | *ZNF767P* |
| Green | 726 | UP | 6.5E-04 | End, Per | Development, Angiogenesis | *COL1A2* |
| Midnightblue | 39 | DOWN | 6.5E-04 | Ast | - | *ENHO* |
| Cyan | 59 | UP | 2.0E-03 | - | - | *BTBD19* |
| Yellow | 848 | DOWN | 3.1E-03 | Ast, Oli | Vesicles | *NEBL* |
| Greenyellow | 105 | DOWN | 1.0E-02 | Ex, In | Synaptic signaling | *CNR1* |
| Turquoise | 1,904 | DOWN | 1.2E-02 | Ex, In | Synaptic signaling | *CAMK2B* |
| Tan | 103 | UP | 1.5E-02 | - | Interferon/Immune | *PARP9* |
| Magenta | 244 | - | 0.134 | - | - | *LOC100996573* |
| Salmon | 87 | - | 0.153 | - | Protein folding | *DNAJB1* |
| Brown | 1,043 | - | 0.210 | Oli, Ast, Per | Development, Cytoskeleton | *SORBS1* |
| Pink | 314 | - | 0.210 | Mic | Immune | *NCKAP1L* |

### Pseudotime analysis

The pseudotime trajectory was generated modeling the brain region as the prior factor of interest. As shown in the PCA analysis in **Figure 6A**, the trajectories for AMY and OBT converge from distinct starting points possibly due to brain region heterogeneity. However, these results suggest that the pathology might drive similar changes across the two brain regions, resulting in converging trajectories. Interestingly, we detected a significant difference of pseudotime between cases and controls, with an increase in COVID-19+ cases (t-test: p = 0.0006 in AMY, and p = 0.011 in OBT) (**Figure 6B**). We detected a total of 5,731 (amygdala) and 4,394 (olfactory bulb) genes significantly correlated with pseudotime trajectories (FDR < 0.05). We conducted pathways analysis and detected similar pathways between the two regions. Across positively correlated genes we observed immune system and extracellular matrix pathways in both amygdala and olfactory bulb. Finally, negatively correlated genes were enriched for neuronal and synaptic pathways. The only differential pathways we detected were related to platelet activation processes/hemostasis, which was only detected in the olfactory bulb (**Figure 7**; **Table S6** and **S7**).

**Figure 6.**
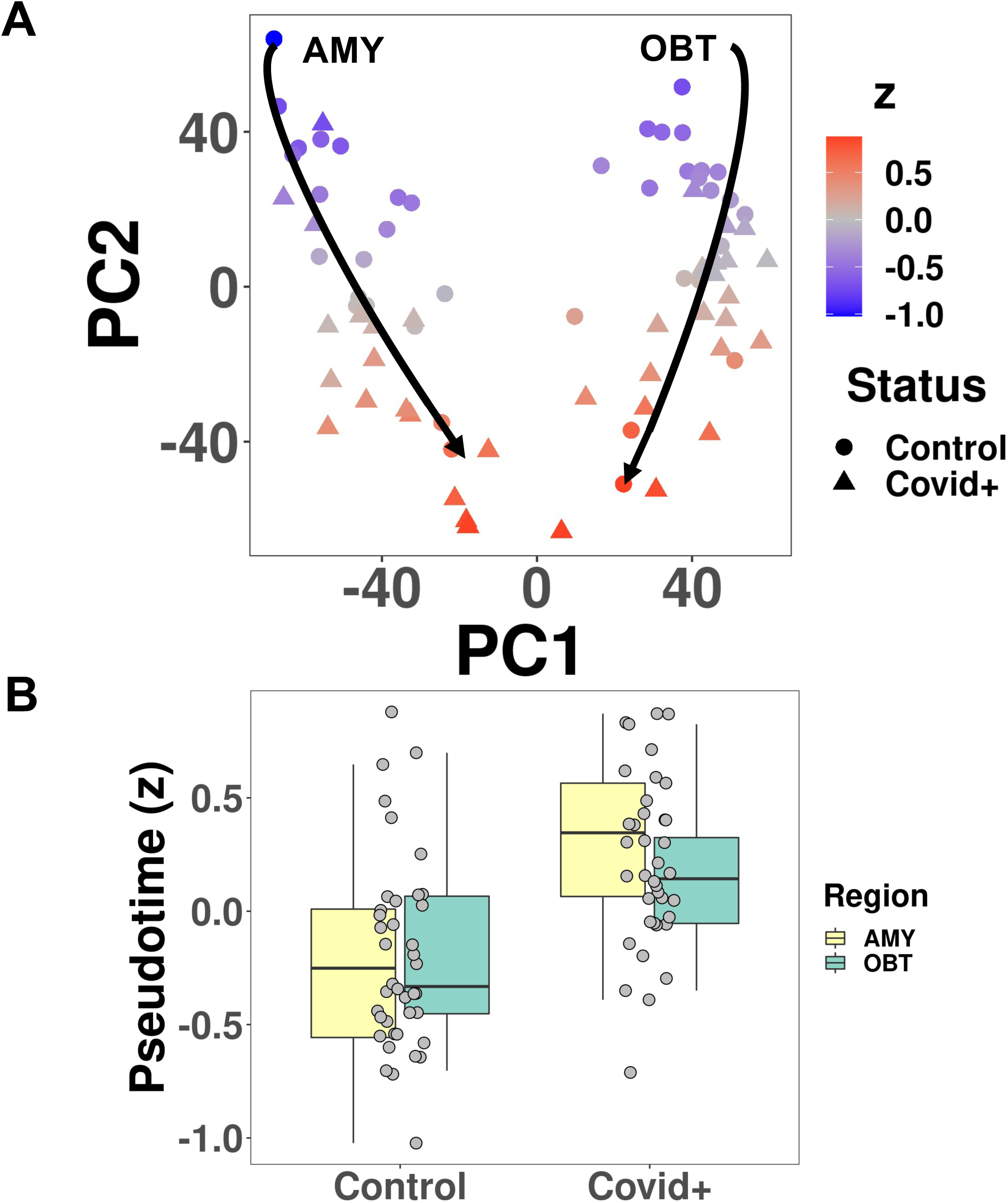
Pseudotime analysis in amygdala and olfactory bulb A)PCA analysis showing converging trajectories for amygdala (left) and olfactory bulb (right). B)Association between pseudotime and disease status, showing an increase of pseudotime in Covid+ cases.

**Figure 7.**
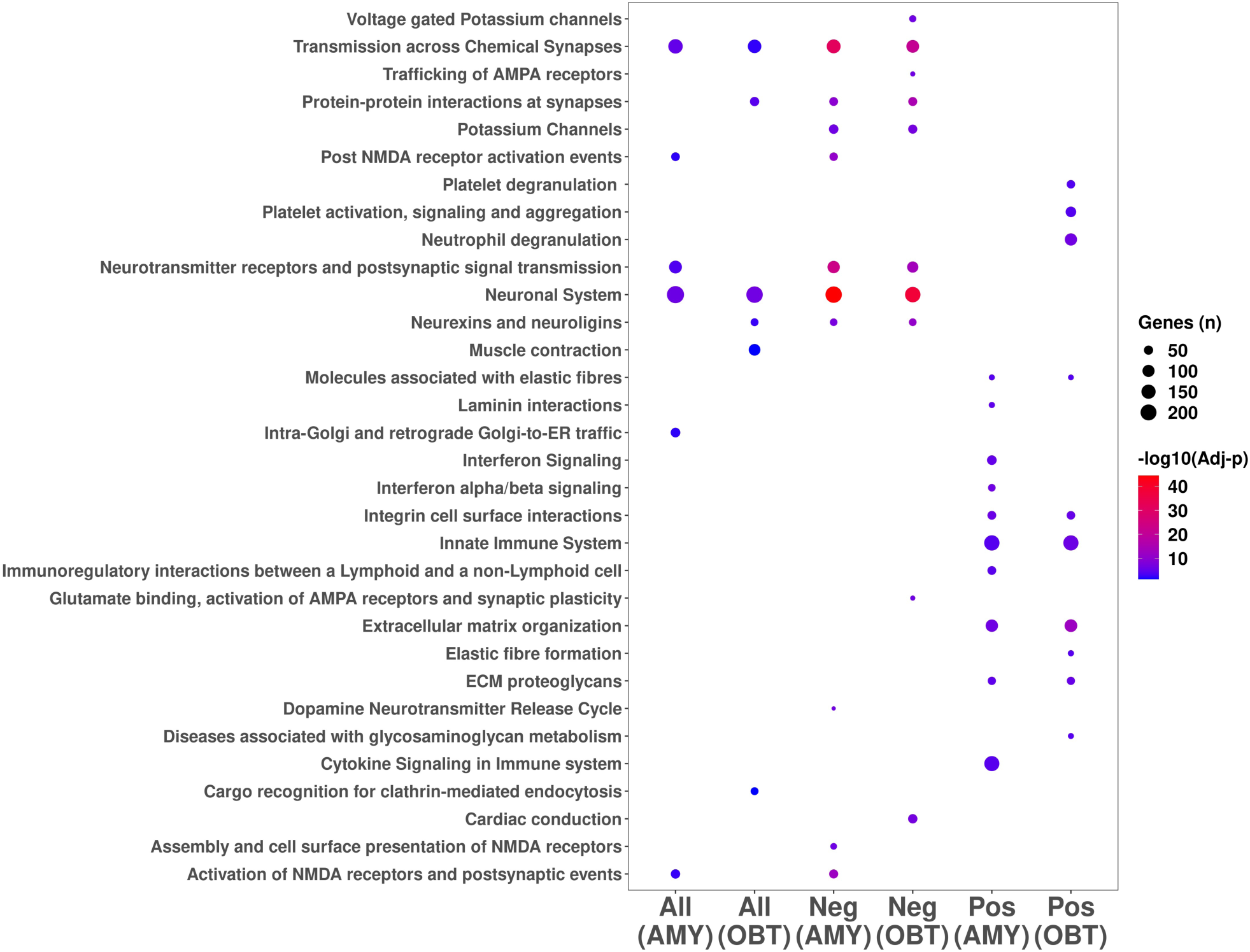
Pathways analysis results (top 10 pathways) conducted using on the genes significantly correlated with pseudotime in amygdala and olfactory bulb. All: all significantly corrected genes Pos: positive correlation Neg: negative correlation

We extracted the genes significantly associated with pseudotime but not differentially expressed between COVID-19+ and controls with the goal of highlighting genes and processes not detectable in the differential expression analysis. We detected a total of 4,275 genes only associated with pseudotime in amygdala and 3,102 only associated with pseudotime in olfactory bulb (**Table S8** and **Table S9**, respectively). The top genes for AMY and OBT are reported in **Figure S3**. The top three genes for amygdala were *SCN3B, BASP1* and *ADD2* (negatively correlated), and the top three genes for the olfactory bulb were *MAP7D2, ADARB1* and *LMTK2*.

## DISCUSSION

In this study we conducted RNA sequencing in postmortem brain tissue from COVID-19 patients and controls, characterizing the amygdala (AMY) and olfactory bulb (OBT). Overall, we detected a higher dysregulation in OBT than AMY, demonstrated by the number of DEGs detected in the two regions (5,405 in OBT vs 1,283 in AMY), and by the results from the WGCNA analysis. Specifically, the coexpression networks detected in AMY were not significantly different between patients and controls, whereas we detected 11 modules with significant changes in OBT. At the pathway level, we detected similar biological processes enriched in both regions, these included the downregulation of synaptic genes and immune system activation, but also specific processes (olfactory/taste pathways and angiogenesis/platelet aggregation in OBT). Pseudotime analysis uncovered converging trajectories highlighting some common processes involved in disease progression which included the immune system response and the downregulation of synaptic/neuronal genes. Finally, we were able to detect genes associated with disease progression not observed in the differential expression analysis.

With a much larger number of DEGs in OBT than in AMY, with several co-expression network changes between patients and controls in OBT, but no significant differences in AMY, the developing picture of OBT as central to CNS COVID-19 is supported by our results. OBT is considered one of the likely entry-points of SARS-CoV-2 in the human brain^54,58,63-65^, and this fact might explain the larger number of transcriptional changes observed. Ageusia and anosmia are typical symptoms of COVID-19 ^64,66,67,115,116^, being a probable consequence of the early viral colonization of the primary olfactory neuroepithelium. We detected a significant enrichment and upregulation of olfactory/taste receptor genes in OBT among the upregulated DEGs and in the red WGCNA module (n = 587 genes), respectively. The hub gene in the coexpression network enriched for taste receptors and signaling was *DMTF1* (Cyclin D Binding Myb Like Transcription Factor 1), encoding for a transcription factor induced by the oncogenic Ras signaling pathways and functioning as a tumor suppressor. These pronounced gene expression changes in OBT might be due to direct OBT neuroinvasion or indirect effects of systemic inflammation, but it also seems possible that this may represent a reaction to structural or functional deafferentation resulting from direct SARS-CoV-2 infection and inflammation of primary olfactory neuroepithelium ^117,118^.

Some of our analyses converge towards an increase of immune system activity in both regions. Immune pathways are upregulated in AMY, and, by WGCNA analysis, we found an upregulated module in the OBT enriched for the same processes. The same results were detected during the pseudotime analysis. Additionally, across upregulated genes we detected a significant enrichment of microglia genes in both regions and astrocyte genes in AMY. The hub gene detected in the immune module was *PARP9* (Poly(ADP-Ribose) Polymerase Family Member 9) which is involved in interferon-mediated antiviral defense ^119,120^. It has been recently demonstrated that the ectopically expressed SARS-CoV-2 Nsp3 macrodomain hydrolyzes PARP9/DTX3L-dependent ADP-ribosylation induced by IFN signaling suggesting a role for this modification as a putative effector of the IFN response ^121^. However, previous reports demonstrated an induction of interferon signaling genes and IFNγ-induced STAT1 phosphorylation ^119,120^. Neuroinflammation has been widely reported in COVID-19 disease ^122-124^ but relatively few reports characterize this in detail and broad confirmation is lacking. Additionally, there are considerable interindividual differences in the circulating immune profile, as well as changes during the clinical course and in relation to the ultimate outcome, with some evidence of an early immunosuppression or maladapted immune response ^125-127^. Early lymphopenia, including decreases in circulating CD4+ and CD8+ cells, are relatively common ^127^. Generally, however, the immune response triggered by COVID-19 infection results in an increase of serum proinflammatory cytokines as IL-1, IL-6, IL-10 and TNF-a ^128,129^. TNF-α can cross the blood-brain-barrier (BBB) or reach the brain via circumventricular organs (CVOs) ^130^ and activate microglia and astrocytes ^131^. This triggers an increase in phagocytosis but also secretion of inflammatory molecules by the microglia including glutamate and quinolinic acid ^132^, resulting in glutamate release and upregulation of NMDA receptors, possibly inducing altered learning, memory and neuroplasticity ^131^. All this evidence supports how neuroinflammation may contribute to the cognitive symptoms reported in COVID-19 disease ^129^.

We detected a deregulation of synaptic and neuronal genes in both OBT and AMY associated with COVID-19 progression, as demonstrated by pathways, WGCNA and pseudotime analysis. Some of the relevant genes were the hubs of the two distinct WGCNA coexpression networks deregulated in OBT, enriched for synaptic and neuronal processes as well as for both excitatory and inhibitory neuronal genes. *CNR1* (cannabinoid receptor 1 – CB1) is the hub of the greenyellow module, and mediates the biological activity of both endogenous and exogenous cannabinoids centered on psychoactive functions ^133^. There are multiple lines of evidence regarding the effects of cannabinoid receptors on viral infection. Activation of CB1 inhibits the production of pro-inflammatory mediators, such as NO or TNFα, and inhibit Ca2+ channels. On one side, the activation of CB1 increases the progression of viral diseases through an immunosuppressive effect, but on the other side it might inhibit the immune effects derived from the viral infection by eliciting and immunoprotective profile ^133^. In our study we see a deregulation of the network regulated by CB1, and we hypothesize this might be a feedback effect aimed at reducing the neuroinflammatory state induced by the viral infection. The hub gene of the turquoise module (n = 1,904 genes) is *CAMK2B* (Calcium/Calmodulin Dependent Protein Kinase II Beta), and is involved in dendritic spine and synapse formation, neuronal plasticity and development ^134^, specifically during cytoskeleton reorganization. Additionally, *CAMK2B* mutations and mRNA alterations were associated with neurodevelopmental diseases, epilepsy and intellectual disability and other neuropsychiatric diseases such as schizophrenia ^135-137^. Cognitive and attention deficits, new-onset anxiety, depression, psychosis and seizures have been observed in some patients with COVID-19, unrelated to respiratory insufficiencies, suggesting brain related alterations that are independent of oxygen saturation ^138^. These symptoms, but also excitotoxicity and synaptic/neuronal loss may be the consequence of the cytokine storm induced by COVID-19 infection ^130-132^. We hypothesize that deregulation of the synaptic genes observed in the *CAMK2B* network might be the consequence of immune system activation effects caused by COVID-19 infection and that these changes may be the critical ones responsible for the short and long-term neurological effects. CAMK2B might be an interesting candidate target to reverse and minimize these short and long term COVID-19 neurological effects. In summary, the results for the WGNCA “synaptic” modules highlight two differential biological processes: a feedback effect of the cannabinoid system to limit the damages of neuroinflammation and synaptic disfunctions driven by the neuroinflammation induced by the infection. It is important to stress that the two modules are not related (**see Table S2**) representing two distinct biological processes.

One of the OBT-specific processes we identified was angiogenesis/blood vessel development (WGCNA – green module) and platelet activation/hemostasis (pseudotime analysis) with upregulation of these processes in COVID-19+ patients. Additionally, we detected enrichment of endothelial cell and pericyte specific genes in OBT. The hub gene in the green module was *COL1A2 (*Collagen Type I Alpha 2 Chain), encoding for the α2 polypeptide of the Type I collagen, the most abundant collagen type in the human body and also a powerful angiogenesis inducer ^139^. SARs-CoV-2 has been hypothesized to enter brain endothelial cells activating neutrophils, macrophages, and thrombin production, promoting microthrombi deposition, capillary congestion and ischemic lesions causing tissue hypoxia and disturbing neurotransmitter synthesis ^49,140^. Angiogenesis is a response against these damages and local hypoxia, and in COVID-19 has been observed in different tissues, including the brain ^141^. *COL1A2* might potentially be a candidate target to reverse/minimize the microvascular damages induced by COVID-19 neuroinflammation.

## CONCLUSIONS

We were able to identify molecular processes and genes associated with the effects of COVID-19 on the brain. Our results indicate the involvement of neuroinflammation as a major driver of COVID-19 effects in the brain, finding an upregulation in OBT and AMY, mostly associated with IFN pathways activation. The effect of *PARP9* as a key driver of neuroinflammation should be further investigated, since there are contrasting results from studies investigating the role as an effector of the IFN cascade. Common observed consequences of the neuroinflammation in COVID-19 are both neurological effects and microvascular damages. Accordingly, we identified a coexpression network downregulated in OBT in COVID-19 patients, and more generally a downregulation of synaptic and neuronal genes in both regions. We hypothesize this deregulation might be associated with the short- and long-term neurological effects of the disease. In this context, *CAMK2B* might be a key gene and a candidate target to reverse or slow these neurological effects. Finally, we found enrichment for angiogenesis and platelet activation genes, where the angiogenesis gene *COL1A2* might be a potential candidate target to antagonize the vascular COVID-19 consequences. All of these results stand in need of further investigation both *in vivo* and *in vitro* to assess their potential role in SARS-CoV-2 infection, response, and COVID-19 development and short-/long-term sequelae.

## Data Availability

The RNA sequencing datasets generated during the current study are available from the corresponding author on reasonable request.

## ACKNOWLEDGEMENTS

This project was supported by a COVID-19 Supplement to a National Institute on Aging grant, (3P30AG019610-20S1), submitted in response to a Notice of Special Interest (NOSI) issued by the National Institute on Aging (NOT-AG-20-022). Biospecimens from the Banner Sun Health Research Institute Brain and Body Donation Program, including those presented in this report, are available to qualified researchers upon request from https://www.brainandbodydonationregistration.org/.

Some of the results published here are in part based on data obtained from the AD Knowledge Portal (https://adknowledgeportal.org). Study data were provided by the Rush Alzheimer’s Disease Center, Rush University Medical Center, Chicago. Data collection was supported through funding by NIA grants P30AG10161 (ROS), R01AG15819 (ROSMAP; genomics and RNAseq), R01AG17917 (MAP), R01AG30146, R01AG36042 (5hC methylation, ATACseq), RC2AG036547 (H3K9Ac), R01AG36836 (RNAseq), R01AG48015 (monocyte RNAseq) RF1AG57473 (single nucleus RNAseq), U01AG32984 (genomic and whole exome sequencing), U01AG46152 (ROSMAP AMP-AD, targeted proteomics), U01AG46161(TMT proteomics), U01AG61356 (whole genome sequencing, targeted proteomics, ROSMAP AMP-AD), the Illinois Department of Public Health (ROSMAP), and the Translational Genomics Research Institute (genomic).

