## Supplementary material for "Olfactory Bulb and Amygdala Gene Expression Changes in Subjects Dying with COVID-19": Figure S1

A B

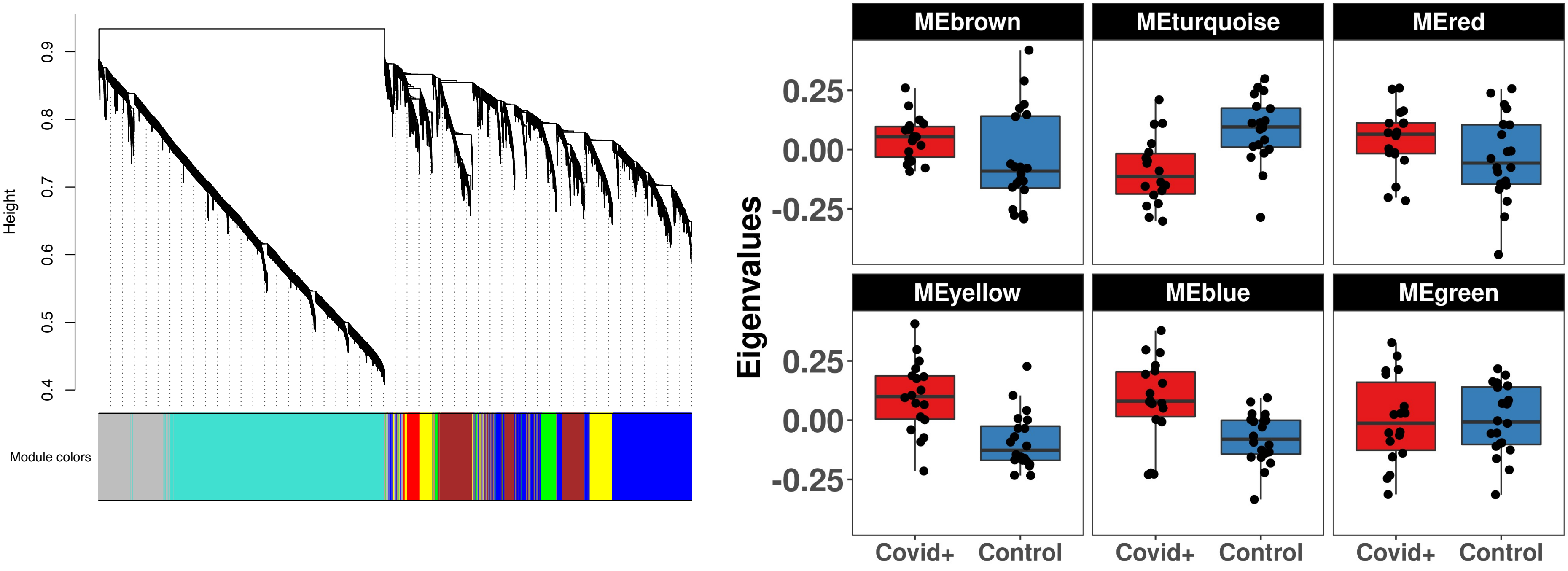


C

| ProbeID | Genes (n) | logFC | AveExpr | p | adj-p |
|---|---|---|---|---|---|
| Brown | 1100 | 0.098 | -5.43E-18 | 0.068 | 0.400 |
| Turquoise | 3049 | -0.065 | 1.43E-19 | 0.226 | 0.400 |
| Red | 204 | 0.061 | -1.07E-17 | 0.252 | 0.400 |
| Yellow | 701 | 0.059 | 2.53E-18 | 0.267 | 0.400 |
| Blue | 1790 | -0.020 | -1.88E-17 | 0.708 | 0.849 |
| Green | 291 | 0.002 | 3.25E-18 | 0.974 | 0.974 |

**Figure S1**. WGCNA analysis in amygdala. We detected 6 coexpression modules (A) not differentially expressed between Covid+ and Controls (B and C)
