## Supplementary figures and images for "Olfactory Bulb and Amygdala Gene Expression Changes in Subjects Dying with COVID-19"

### Figure S2

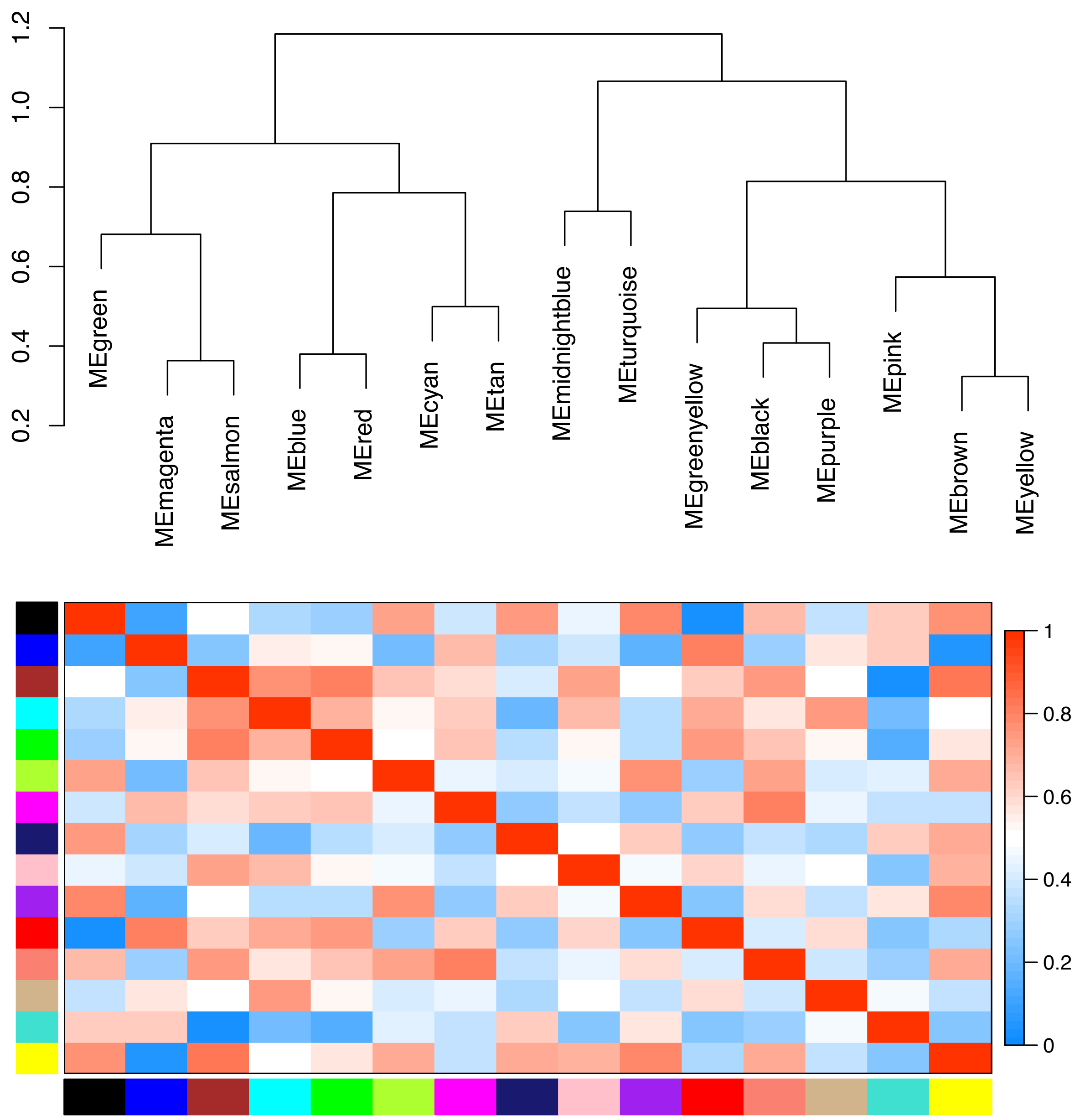


**Figure S2**. Dendrogram representing the relationship between WGCNA modules from OBT
