## Supplementary material for "Olfactory Bulb and Amygdala Gene Expression Changes in Subjects Dying with COVID-19": Figure S3

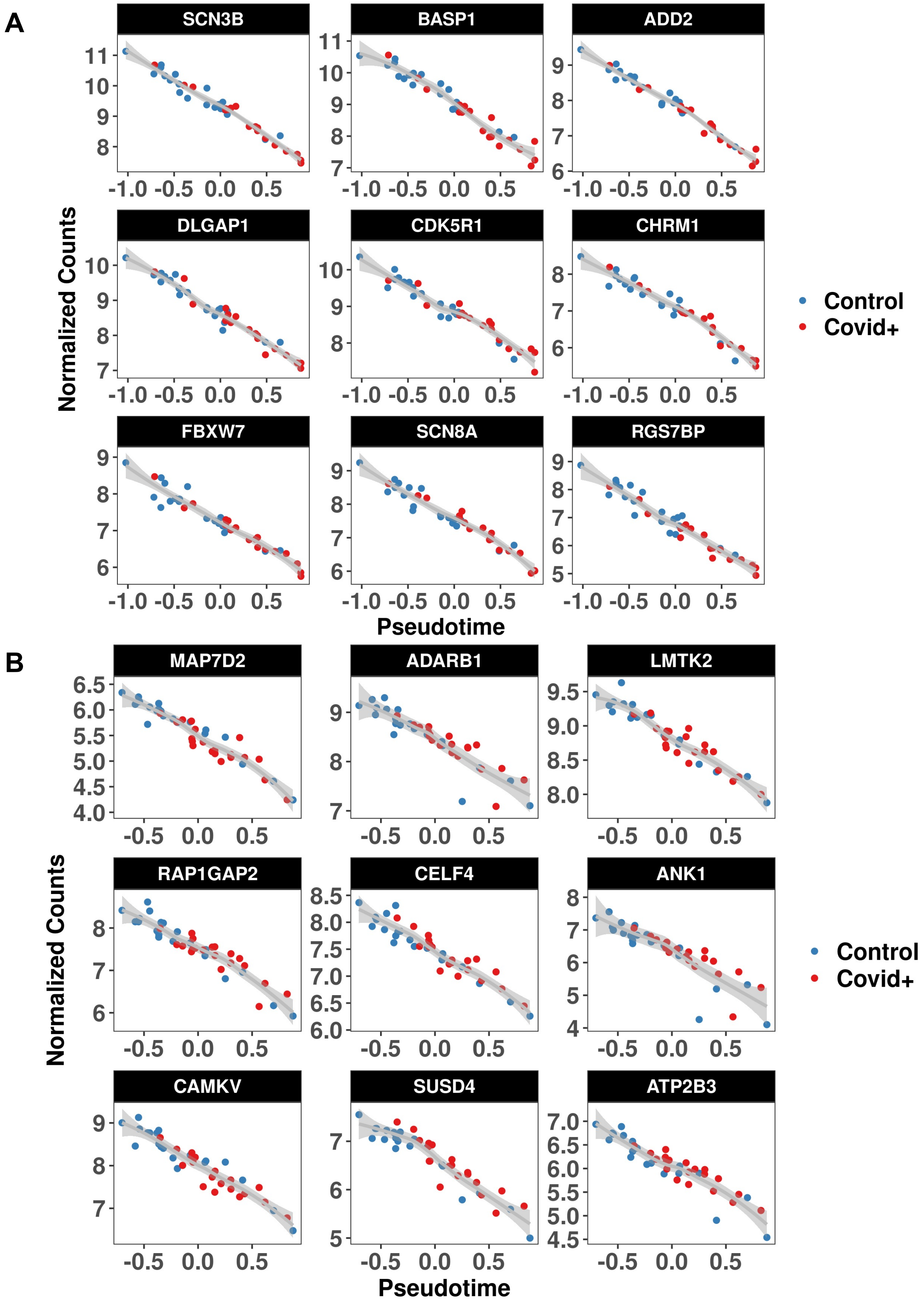


**Figure S3**. Top nine genes significantly correlated with pseudotime in amygdala (A) and olfactory bulb (B) but not differentially expressed between Covid+ and Controls (FDR > 0.05)
